# CHILDHOOD DEATHS, DEPRIVATION, AND MODIFIABLE FACTORS: FINDINGS FROM THE NATIONAL CHILD MORTALITY DATABASE

**DOI:** 10.1101/2022.07.04.22276688

**Authors:** David Odd, Sylvia Stoianova, Tom Williams, Dawn Odd, Jennifer J Kurinczuk, Ingrid Wolfe, Karen Luyt

## Abstract

**Objectives:** The aim of this analysis is to identify and report the patterns of social deprivation in relation to childhood mortality; and identify potential points where public health, social and education interventions or health policy may be best targeted.

**Design:** Decile of deprivation and underlying population distribution was derived using Office for National Statistics (ONS) data. The risk of death was then derived using a Poisson regression model, calculating the increasing risk of death for each increasing deprivation decile.

**Setting:** England

**Participants:** 2688 childhood deaths in England reviewed between the April 2019 and March 2020.

**Main Outcome Measures:** The relationship between deprivation and risk of death; for deaths with, and without modifiable factors.

**Results:** There was evidence of increasing mortality risk with increase in deprivation decile (RR 1.08 (1.07 to 1.10)), with the gradient of risk stronger in children who died with modifiable factors than those without (RR 1.12 (1.09 to 1.15)) vs (RR 1.07 (1.05 to 1.08)). Deprivation sub-domains of Employment, Adult Education, Barriers to Housing and Services, and Indoor Living Environments appeared to be the most important predictors of child mortality

**Conclusions:** There is a clear gradient of increasing child mortality across England as measures of deprivation increase; with a striking finding that this varied little by area, age or other demographic factor. Over a fifth of all child deaths may be avoided if the most deprived half of the population had the same mortality as the least deprived. Children dying in more deprived areas may have a greater proportion of avoidable deaths. Adult employment and education, and improvements to housing, may be the most efficient place to target resources to reduce these inequalities.

## BACKGROUND

The death of every child is a devastating loss that profoundly affects bereaved parents as well as siblings, grandparents, extended family members, friends and professionals. The evidence relating to social deprivation and death is strongest for infant mortality however the effects appear measurable across the life course.[1] A systematic review examining the relationship between social factors and early childhood health and developmental outcomes provides strong evidence that factors such as neighbourhood deprivation, lower parental income, unemployment and educational attainment, lower occupational social class, heavy physical occupational demands, lack of housing tenure, and material deprivation in the household are all independently associated with a wide range of adverse health outcomes.[2]

We know that early child development plays a major role in affecting future life chances and health throughout the life course[3] with adverse exposures having greater impacts on younger children[4]. While initiatives have been proposed to reduce the impact of deprivation on health[5]; babies, children, and young people remain the most vulnerable in society. Currently England has one of the highest infant mortality rates in Europe[6,7] and while much of the variation may be due to socioeconomic factors[8], it is clear that since infant mortality among the most deprived groups continues to rise[9], effective policies and other interventions are either lacking or have not been successfully implemented. While the COVID pandemic continues to impact delivery of social and healthcare programs across the world, the longer term impact on economies and social and healthcare budgets is likely to be substantial, and social inequalities even in developed nations, may worsen.

The National Child Mortality Database (NCMD) Programme was established in 2018 to collate and analyse data about all children in England who die before their 18^th^ birthday, with statutory death notifications required within 48 hours[10]. The data are collated from the 58 Child Death Overview Panels (CDOPs) in England who carry out detailed analysis of the circumstances of death and identify the modifiable contributory factors relevant to the death as part of the child death review (CDR) process with the aim of identifying common themes to guide learning and inform actions to reduce future child deaths.[11] The CDR process is statutory, with the Children Act 2004 mandating the review and analysis of all child deaths so the circumstances of death that relate to the welfare of children locally and nationally, or to public health and safety, are identified and understood, and preventive actions established. This work is based on the NCMD Programme’s first thematic report[12].

### Aims

The aim of this analysis is to identify and report the patterns of social deprivation, and modifiable factors in relation to childhood mortality, and identify potential intervention points and high risk groups where public health, social and education, or health policy may be best targeted.

## METHODS

Three external sources of data were linked to the child death review data using the smallest geographical level of the deprivation index (the Lower Super Output Area (LSOA)). This allowed further estimation of the population estimates of age and sex[13], its rural (Rural town and fringe, Rural village) or urban (Urban city and town, Urban major conurbation) status[14] and its location in England (East Midlands, East of England, London, North East, North West, South East, South West, West Midlands, Yorkshire and the Humber)[15]

### Exploratory Variables

For the primary exploratory analysis variables of interest were:

- Age of death (age as a continuous measure) then coded for analysis and presentation as <1 year, 1-4 years, 5-9 years, 10-14 years and 15-17 years).
- Sex (male, female, or missing (including “indeterminate”, “not known”, “N/A”, “NULL” etc)).
- Area of residence: Urban vs Rural[15]
- Region of England.
- Ethnicity was coded as White, Asian or British Asian, Black or British Black, Mixed or Other.

### Specific Detailed Data from Child Death Review Process

The CDOP is responsible for identifying any modifiable factors in relation to the child’s death. Modifiable factors are those which may have contributed to the death of the child and which might, by means of a locally or nationally achievable intervention, be modified to reduce the risk of future deaths. Factors identified by the CDOP were further classified as:

- Characteristics of the child (e.g. loss of key relationships, risk taking behaviour, comorbidity, prematurity, congenital anomaly, learning disability, eating disorder, suicidal ideation or previous suicide attempt)
- Social Environment (e.g. abuse, parenting, consanguinity, financial pressures/hardship)
- Physical Environment (e.g. animal attack, homicide, vehicle related deaths, safety within the home, unsafe infant sleeping practices, and public equipment)
- Service Provision (e.g. gaps in service provision, failure to follow guidelines, poor communication, staffing issues and bed occupancy)

Category of death was allocated by the CDOP while reviewing the case and was categorised as; Acute Medical and Surgical, Congenital Anomalies, Chronic Medical, Deliberately inflicted injury, Infection, Malignancy, Perinatal, Sudden Unexplained Death in Childhood (SUDIC), Suicide or deliberate self-inflicted harm or Trauma.

### Analysis

Initially the characteristics of all child deaths reviewed between April 2019 and March 2020 were derived, stratified by the available covariates (listed above). Next we derived the proportion of deaths in each deprivation decile. Evidence of any trend in proportions by increasing deprivation decile were tested using a nonparametric test for trend across ordered groups[16]. This was then repeated for each category of death.

Second, to assess any association between deprivation and the risk of death, the population distribution was derived using ONS data for each LSOA producing a dataset with the predicted numbers of children of each age, sex, rural/urban status and region. The risk of death was then derived using a Poisson regression model, calculating the increasing risk of death for each increasing deprivation decile, with the model then adjusted for the other known underlying population characteristics or possible confounders (sex, age, rural/urban area and region). Lastly both the unadjusted and adjusted model were repeated for each reported category of death and tested (using the likelihood ratio test) to assess if the association between deprivation measures and overall mortality was modified by sex, age category, region or rural/urban status. Finally for overall mortality a separate model was derived for those children in the lowest five vs the highest five deciles of deprivation, and used to estimate the population attributable risk fraction for those children living the in the most deprived five deciles.

Next, to interrogate the possible causes we initially derived the number, proportion and evidence of trend[16] of modifiable factors identified at the CDOP review across each deprivation decile. We then calculated the increasing risk of death for each increasing deprivation decile separately for those deaths with, or without, modifiable factors identified. The analyses were repeated, stratified by the sub-categories of modifiable factors, and by the category of death.

### Patient and public involvement

Parent and public involvement is at the heart of the NCMD programme. We are indebted to Charlotte Bevan (Sands - Stillbirth and Neonatal Death Charity), Therese McAlorum (Child Bereavement UK) and Jenny Ward (Lullaby Trust), who represent bereaved families on the NCMD programme steering group.

## RESULTS

A total of 2688 childhood deaths were reviewed by CDOPs between April 2019 and March 2020 and linked to deprivation measures (Table 1). The most common age at death was less than 1 year (62.3%) and more boys than girls died (56.5 vs 43.6% respectively). The majority lived in areas defined as urban (87.8%) and most were of a white ethnic background (65.0%). Deaths were more common in children in the most deprived deciles (Table 2) (p=0.003). When looking at the categories of death, deaths due to acute medical or surgical disease (p=0.017), congenital anomalies (p=0.003), chronic medical (p=0.006), deliberate inflicted injury (p=0.025), infection (p=0.021), perinatal (p=0.006), SUDIC (p=0.003) and trauma (p=0.038) appeared to be associated with increasing deprivation. There was little evidence of an association between increasing deprivation and deaths from malignancy (p=0.326) or suicide or deliberate self-inflicted harm (p=0.296).

**Table 1.** Characteristics of the populations of child deaths reviewed by CDOPs in England during 2019/2020.

| **Measure** | **N** | **Child deaths reviewed 2019/2020** |
| --- | --- | --- |
| **All Deaths** | 2688 | - |
| **Age of Death** | 2688 |  |
| <1 year |  | 1675 (62.3%) |
| 1-4 Years |  | 322 (12.0%) |
| 5-9 Years |  | 211 (7.9%) |
| 10-14 Years |  | 227 (8.4%) |
| 15-17 Years |  | 253 (9.4%) |
| **Sex** | 2670 |  |
| Male |  | 1505 (56.4%) |
| Female |  | 1165 (43.6%) |
| **Area of residence** | 2688 |  |
| Rural |  | 328 (12.2%) |
| Urban |  | 2360 (87.8%) |
| **Ethnicity** | 2390 |  |
| White |  | 1554 (65.0%) |
| Asian or British Asian |  | 427 (17.9%) |
| Black or British Black |  | 188 (7.9%) |
| Mixed |  | 136 (5.7%) |
| Other |  | 85 (3.6%) |
| **Region of residence** | 2688 |  |
| East Midlands |  | 214 (8.0%) |
| East of England |  | 211 (8.2%) |
| London |  | 473 (17.6%) |
| North East |  | 109 (4.1%) |
| North West |  | 362 (13.5%) |
| South East |  | 336 (12.5%) |
| South West |  | 232 (8.6%) |
| West Midlands |  | 400 (12.9%) |
| Yorkshire and the Humber |  | 341 (12.7%) |

**Table 2.**
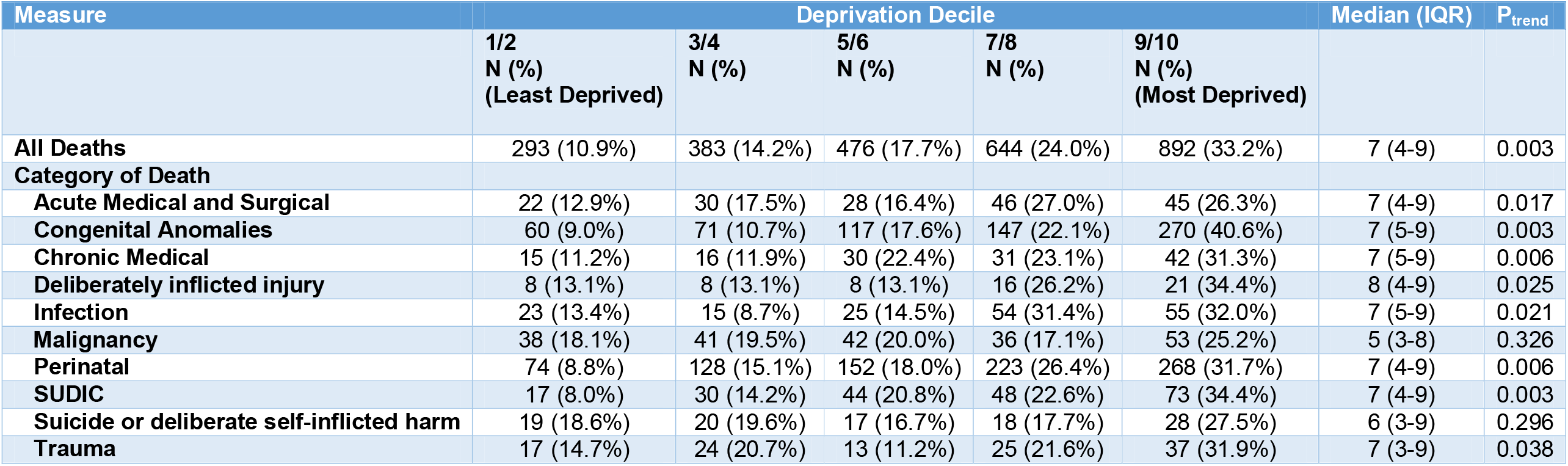
Deaths by deprivation decile, stratified by the category of death and patient characteristics of child deaths.

| Measure | Deprivation Decile |  |  |  |  | Median (IQR) | P<sub>trend</sub> |
| --- | --- | --- | --- | --- | --- | --- | --- |
|  | 1/2<br>N (%)<br>(Least Deprived) | 3/4<br>N (%) | 5/6<br>N (%) | 7/8<br>N (%) | 9/10<br>N (%)<br>(Most Deprived) |  |  |
| **All Deaths** | 293 (10.9%) | 383 (14.2%) | 476 (17.7%) | 644 (24.0%) | 892 (33.2%) | 7 (4-9) | 0.003 |
| **Category of Death** |  |  |  |  |  |  |  |
| **Acute Medical and Surgical** | 22 (12.9%) | 30 (17.5%) | 28 (16.4%) | 46 (27.0%) | 45 (26.3%) | 7 (4-9) | 0.017 |
| **Congenital Anomalies** | 60 (9.0%) | 71 (10.7%) | 117 (17.6%) | 147 (22.1%) | 270 (40.6%) | 7 (5-9) | 0.003 |
| **Chronic Medical** | 15 (11.2%) | 16 (11.9%) | 30 (22.4%) | 31 (23.1%) | 42 (31.3%) | 7 (5-9) | 0.006 |
| **Deliberately inflicted injury** | 8 (13.1%) | 8 (13.1%) | 8 (13.1%) | 16 (26.2%) | 21 (34.4%) | 8 (4-9) | 0.025 |
| **Infection** | 23 (13.4%) | 15 (8.7%) | 25 (14.5%) | 54 (31.4%) | 55 (32.0%) | 7 (5-9) | 0.021 |
| **Malignancy** | 38 (18.1%) | 41 (19.5%) | 42 (20.0%) | 36 (17.1%) | 53 (25.2%) | 5 (3-8) | 0.326 |
| **Perinatal** | 74 (8.8%) | 128 (15.1%) | 152 (18.0%) | 223 (26.4%) | 268 (31.7%) | 7 (4-9) | 0.006 |
| **SUDIC** | 17 (8.0%) | 30 (14.2%) | 44 (20.8%) | 48 (22.6%) | 73 (34.4%) | 7 (4-9) | 0.003 |
| **Suicide or deliberate self-inflicted harm** | 19 (18.6%) | 20 (19.6%) | 17 (16.7%) | 18 (17.7%) | 28 (27.5%) | 6 (3-9) | 0.296 |
| **Trauma** | 17 (14.7%) | 24 (20.7%) | 13 (11.2%) | 25 (21.6%) | 37 (31.9%) | 7 (3-9) | 0.038 |

When estimating the relative risk of death using an unadjusted Poisson model, there was an increasing risk of all cause mortality as measures of deprivation increased (RR 1.11 (95% CI 1.09-1.12), p<0.001); but also for death categorised as acute medical or surgical (p=0.030), congenital anomalies (p<0.001), chronic medical (p=0.004), deliberately inflicted injury (p=0.009), infection (p<0.001), perinatal (p<0.001), and SUDIC (p<0.001) (Table 3). After adjusting for age, sex, region and rural status, the association with all cause mortality (RR 1.08 (95% CI 1.07-1.10), p<0.001) and for congenital anomalies (p<0.001), chronic medical (p=0.007), deliberately inflicted injury (p=0.040), infection (p<0.001), perinatal (p<0.001), and SUDIC (p<0.001) remained. However, in the adjusted analysis, the association between death in the acute medical or surgical category with increasing measures of deprivation weakened slightly (p=0.052). There was little evidence to suggest an association with malignancy (p=0.868), suicide or deliberate self-inflicted harm (p=0.831) or trauma (p=0.075) in the unadjusted (p=0.868, p=0.831 and p=0.075 respectively) or in the adjusted analyses (p=0.979, p=0.475 and p=0.174 respectively) (Table 3).

**Table 3.** Relative risk of death for increasing deprivation stratified by category of death, and testing for interactions by characteristics of the child deaths.

| Measure | Unadjusted |  |  |  | Adjusted* |  |  |  |
| --- | --- | --- | --- | --- | --- | --- | --- | --- |
|  | n | RR 95% CI | p |  | n | RR 95% CI | p |  |
| **All Deaths** | 2688 | 1.11 (1.09-1.12) | <0.001 |  | 2670 | 1.08 (1.07-1.10) | <0.001 |  |
| **Acute Medical and Surgical** | 171 | 1.06 (1.01-1.12) | 0.030 |  | 170 | 1.06 (1.00-1.12) | 0.052 |  |
| **Congenital Anomalies** | 665 | 1.17 (1.14-1.21) | <0.001 |  | 658 | 1.13 (1.10-1.17) | <0.001 |  |
| **Chronic Medical** | 134 | 1.09 (1.03-1.16) | 0.004 |  | 134 | 1.09 (1.02-1.17) | 0.007 |  |
| **Deliberately inflicted injury** | 61 | 1.13 (1.03-1.24) | 0.009 |  | 61 | 1.11 (1.00-1.22) | 0.040 |  |
| **Infection** | 172 | 1.13 (1.07-1.19) | <0.001 |  | 172 | 1.11 (1.05-1.18) | <0.001 |  |
| **Malignancy** | 210 | 1.00 (0.95-1.04) | 0.868 |  | 210 | 1.00 (0.95-1.05) | 0.979 |  |
| **Perinatal** | 845 | 1.11 (1.09-1.14) | <0.001 |  | 836 | 1.07 (1.04-1.10) | <0.001 |  |
| **SUDIC** | 212 | 1.13 (1.08-1.19) | <0.001 |  | 211 | 1.10 (1.05-1.16) | <0.001 |  |
| **Suicide or deliberate self-inflicted harm** | 102 | 1.01 (0.94-1.08) | 0.831 |  | 102 | 1.03 (0.96-1.10) | 0.475 |  |
| **Trauma and other external factors** | 116 | 1.06 (0.99-1.13) | 0.075 |  | 116 | 1.05 (0.98-1.12) | 0.174 |  |
| **Interactions** |  | **RR 95% CI** | **p** | **p<sub>interaction</sub>** |  | **RR 95% CI** | **p** | **p<sub>interaction</sub>** |
| **Sex** |  |  |  | 0.227 |  |  |  | 0.196 |
| **Female** | 1165 | 1.11 (1.09-1.13) | <0.001 |  | 1165 | 1.07 (1.05-1.09) | <0.001 |  |
| **Male** | 1505 | 1.10 (1.08-1.11) | <0.001 |  | 1505 | 1.09 (1.07-1.11) | <0.001 |  |
| **Age** |  |  |  | 0.003 |  |  |  | <0.001 |
| **<1 year** | 1675 | 1.11 (1.09-1.13) | <0.001 |  | 1659 | 1.10 (1.08-1.12) | <0.001 |  |
| **1-4 Years** | 322 | 1.10 (1.06-1.14) | <0.001 |  | 321 | 1.09 (1.05-1.13) | <0.001 |  |
| **5-9 Years** | 211 | 1.00 (0.96-1.05) | 0.956 |  | 210 | 0.99 (0.95-1.04) | 0.785 |  |
| **10-14 Years** | 227 | 1.07 (1.03-1.12) | 0.002 |  | 227 | 1.07 (1.02-1.11) | 0.006 |  |
| **15-17 Years** | 253 | 1.06 (1.01-1.10) | 0.011 |  | 253 | 1.05 (1.01-1.09) | 0.028 |  |
| **Area** |  |  |  | 0.616 |  |  |  | 0.463 |
| **Urban** | 2360 | 1.10 (1.09-1.12) | <0.001 |  | 2342 | 1.08 (1.06-1.10) | <0.001 |  |
| **Rural** | 328 | 1.12 (1.07-1.17) | <0.001 |  | 328 | 1.10 (1.05-1.16) | <0.001 |  |
| **Region** |  |  |  | 0.074 |  |  |  | 0.165 |
| **East Midlands** | 214 | 1.07 (1.02-1.12) | 0.004 |  | 214 | 1.06 (1.01-1.11) | 0.023 |  |
| **East of England** | 221 | 1.07 (1.02-1.13) | 0.005 |  | 220 | 1.06 (1.01-1.11) | 0.030 |  |
| **London** | 473 | 1.06 (1.02-1.10) | 0.003 |  | 464 | 1.06 (1.01-1.10) | 0.007 |  |
| **North East** | 109 | 1.06 (0.99-1.13) | 0.098 |  | 109 | 1.04 (0.97-1.12) | 0.233 |  |
| **North West** | 362 | 1.10 (1.06-1.14) | <0.001 |  | 360 | 1.08 (1.04-1.12) | <0.001 |  |
| **South East** | 336 | 1.11 (1.07-1.15) | <0.001 |  | 336 | 1.09 (1.05-1.14) | <0.001 |  |
| **South West** | 232 | 1.10 (1.05-1.16) | <0.001 |  | 232 | 1.09 (1.03-1.14) | 0.001 |  |
| **West Midlands** | 400 | 1.16 (1.11-1.20) | <0.001 |  | 395 | 1.14 (1.09-1.19) | <0.001 |  |
| **Yorkshire and the Humber** | 3411 | 1.10 (1.06-1.14) | <0.001 |  | 340 | 1.09 (1.05-1.13) | <0.001 |  |

\* Adjusted for age, sex, region and rural/urban area

There was strong evidence that the association between number of deaths and the deprivation index was modified by age (fully adjusted; p<0.001), but not sex (fully adjusted; p=0.196) or rural/urban status (fully adjusted; p=0.463). In the unadjusted model there was some weak evidence that the relationship may be modified by the region of England (p=0.0743), although this weakened in the adjusted model further (p=0.165).

In the final, adjusted, regression model, estimating the risk of death (adjusted for age, sex and rural/urban area), comparing the risk of death in the most deprived five deciles with the least deprived five deciles, gave compatible results to those from the main analysis (RR 1.47 (1.35-1.60), p<0.001), and a population attributable risk fraction of 21.2% (95% CI 16.7%-25.4%).

The absolute number of deaths where modifiable factors were identified increased as measures of deprivation increased (Figure 1), with additional strong evidence that the proportion of deaths with modifiable contributory factors identified at the CDOP review increased with increasing measures of deprivation; with 24.2% of deaths in the least deprived, compared with 35.1% of deaths in the most (p_trend_<0.001) (Table 4). Children who died with modifiable factors showed a stronger gradient with increasing deprivation (RR 1.12 (1.09-1.15)) compared to those who died without (RR 1.07 (1.05-1.08)). Individually, only those modifiable factors relating to social environment appeared to show this gradient (p<0.001), with less evidence (but small numbers) for those factors around the child, services, or their physical environment. When stratifying by the category of death there was evidence that modifiable factors were more commonly identified in deaths in areas or greater deprivation for congenital anomalies (p=0.001), perinatal (p=0.045) and SUDIC (p=0.045) deaths; with corresponding greater relative risks with deprivation compared to deaths without modifiable factors identified (e.g. Relative risk of death from a congenital abnormality with increasing deprivation was 1.11 (1.07-1.15) for deaths without modifiable factors, and 1.27 (1.16-1.40) for those with).

**Figure 1.**
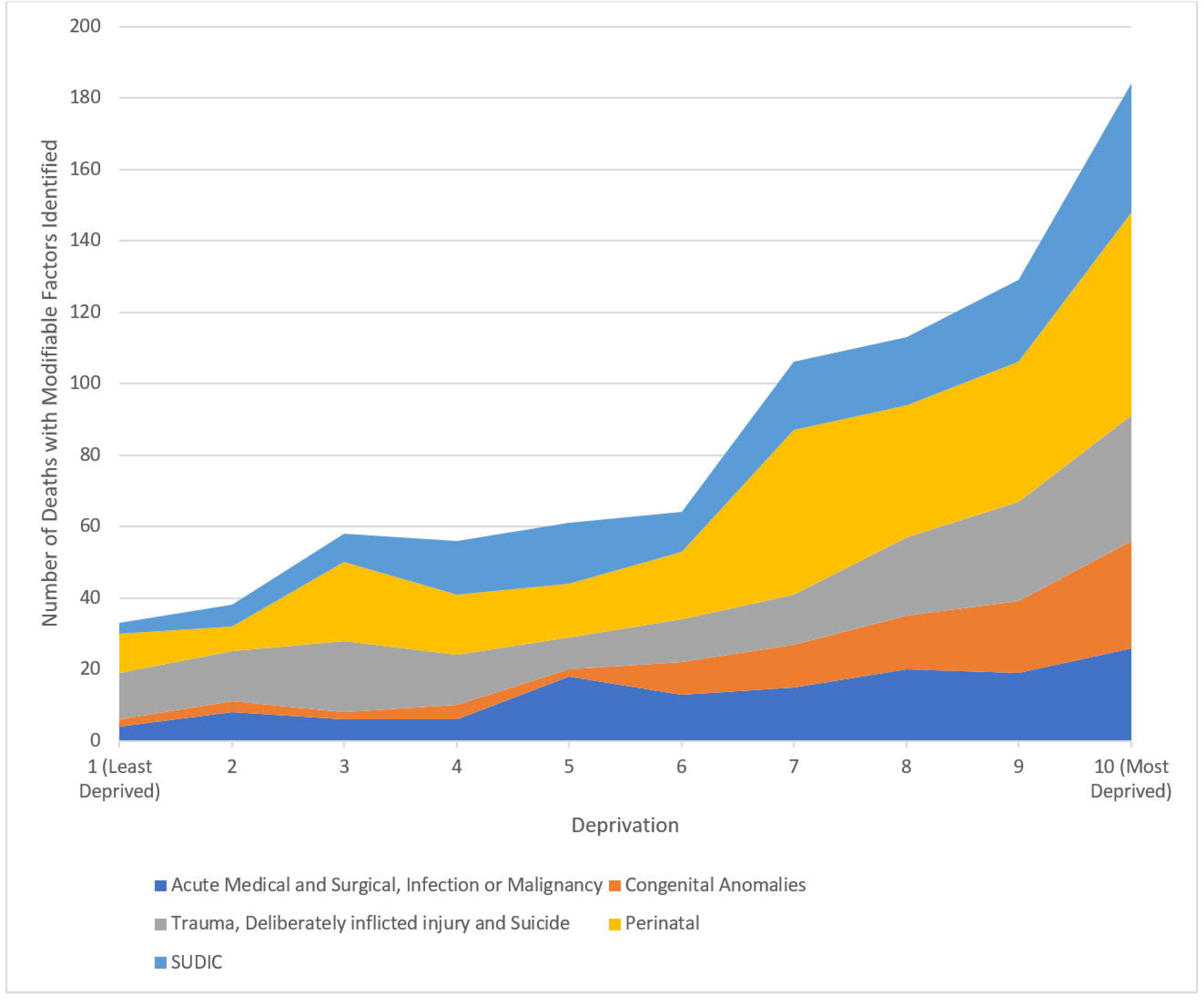
Number of deaths with modifiable factors identified at review, split by measure of local deprivation.

**Table 4.** The number of deaths, in each deprivation decile with identified modifiable factors; and the relative risk of death for each increasing deprivation decile with, or without them; split by category of death.

| Category of Death | Percentage of deaths with modifiable factors |  |  |  |  |  |  | Relative risk of death for increasing deprivation decile* |  |
| --- | --- | --- | --- | --- | --- | --- | --- | --- | --- |
|  | All deciles | Split by Deprivation Decile |  |  |  |  | P<sub>trend</sub> | Death without Modifiable Factors | Deaths with Modifiable Factors |
|  |  | 1/2<br>N (%)<br>(Least Deprived) | 3/4<br>N (%) | 5/6<br>N (%) | 7/8<br>N (%) | 9/10<br>N (%)<br>(Most Deprived) |  |  |  |
| **All Deaths** | 842 (31.3%) | 71 (24.2%) | 114 (29.8%) | 125 (26.3%) | 219 (34.0%) | 313 (35.1%) | <0.001 | 1.07 (1.05-1.08) | 1.12 (1.09-1.15) |
| **Split by type of Modifiable Factors** |  |  |  |  |  |  |  |  |  |
| **Characteristics of the child** | 70 (2.6%) | 9 (3.1%) | 14 (3.7%) | 6 (1.3%) | 15 (2.3%) | 26 (2.9%) | 0.797 | 1.08 (1.07-1.10) | 1.10 (1.01-1.21) |
| **Physical Environment** | 185 (6.9%) | 18 (6.1%) | 30 (7.8%) | 29 (6.1%) | 41 (6.4%) | 67 (7.5%) | 0.764 | 1.08 (1.07-1.10) | 1.08 (1.02-1.14) |
| **Service Provision** | 243 (7.9%) | 26 (8.9%) | 43 (11.2%) | 47 (9.9%) | 57 (8.9%) | 70 (7.9%) | 0.131 | 1.08 (1.07-1.10) | 1.07 (1.02-1.12) |
| **Social Environment** | 416 (15.5%) | 29 (9.9%) | 46 (12.0%) | 51 (10.7%) | 106 (16.5%) | 184 (20.6%) | <0.001 | 1.07 (1.05-1.09) | 1.15 (1.11-1.20) |
| **Split by Category of Death** |  |  |  |  |  |  |  |  |  |
| **Acute Medical and Surgical** | 42 (24.6%) | 5 (22.7%) | 8 (26.7%) | 7 (25.0%) | 9 (20.0%) | 13 (29.0%) | 0.815 | 1.05 (0.98-1.12) | 1.10 (0.98-1.24) |
| **Congenital Anomalies** | 99 (14.9%) | 5 (8.3%) | 6 (8.5%) | 11 (9.4%) | 27 (18.4%) | 50 (18.5%) | 0.001 | 1.11 (1.07-1.15) | 1.27 (1.16-1.40) |
| **Chronic Medical** | 21 (15.7%) | 1 (6.7%) | 2 (12.5%) | 6 (20.0%) | 4 (12.9%) | 8 (19.1%) | 0.597 | 1.09 (1.01-1.17) | 1.14 (0.96-1.35) |
| **Deliberately inflicted injury** | 43 (70.5%) | 4 (50.0%) | 7 (87.5%) | 6 (75.0%) | 12 (75.0%) | 14 (66.7%) | 0.911 | 1.08 (0.90-1.29) | 1.12 (0.99-1.26) |
| **Infection** | 61 (35.5%) | 6 (26.1%) | 1 (6.7%) | 13 (52.0%) | 20 (37.0%) | 21 (38.2%) | 0.126 | 1.07 (1.00-1.15) | 1.20 (1.07-1.33) |
| **Malignancy** | 11 (5.2%) | 0 (0.0%) | 1 (2.4%) | 5 (11.9%) | 2 (5.6%) | 3 (5.7%) | 0.181 | 0.99 (0.94-1.05) | 1.15 (0.91-1.46) |
| **Perinatal** | 270 (32.0%) | 18 (24.3%) | 39 (30.5%) | 34 (22.4%) | 83 (37.2%) | 96 (35.8%) | 0.015 | 1.06 (1.03-1.10) | 1.09 (1.04-1.14) |
| **SUDIC** | 157 (75.1%) | 9 (52.9%) | 23 (76.7%) | 28 (63.6%) | 38 (79.2%) | 59 (80.8%) | 0.045 | 1.02 (0.92-1.12) | 1.14 (1.07-1.21) |
| **Suicide** | 59 (57.8%) | 12 (63.2%) | 9 (45.0%) | 8 (47.1%) | 9 (50.0%) | 21 (75.0%) | 0.317 | 1.01 (0.90-1.12) | 1.04 (0.95-1.14) |
| **Trauma** | 79 (68.1%) | 11 (64.7%) | 18 (75.0%) | 7 (53.9%) | 15 (60.0%) | 28 (75.7%) | 0.743 | 1.00 (0.89-1.12) | 1.07 (0.99-1.17) |

\* Adjusted for age, sex, region and rural/urban area

When analysing the associations between the risk of childhood death and the deprivation sub-domains (Appendix 1), many of the components of the IMD appeared to be closely correlated, with Income and Employment the highest correlation of 0.939 (Appendix 2). The sub-domains selected by the adaptive model, as the strongest associations with childhood deaths (and each categories of death), are shown in Table 5. Measures of deprivation in the domains of Employment, Adult Education, Wider barriers and Indoor Living Environments were identified as most correlated with all cause mortality. Crime also appeared correlated, but in the opposite direction to the others (i.e. increasing measures of deprivation was associated with lower mortality). There was no clear association of any sub-domain and death by malignancy or deliberately inflicted injury; while in contrast the model for perinatal deaths (the single most common category of death) identified measures of Employment and Wider Barriers as possible predictors. Due to the unexpected association between measures of Crime and reductions in risk of death in the adaptive models, a post-hoc analysis was performed to assess the association between this measure and overall mortality. In this model (without the other sub-domain measures of deprivation), increases in measures of deprivation related to crime were associated with increased child mortality (RR 1.06 (1.03-1.09), p<0.001).

**Table 5.** Sub-decile measures identified as stongest associations with childhood death.

| IMD Sub-decile | Category of Death |  |  |  |  |  |  |  |  |  |  |
| --- | --- | --- | --- | --- | --- | --- | --- | --- | --- | --- | --- |
|  | All Deaths | Acute Medical and Surgical | Congenital Anomalies | Chronic Medical | Deliberately inflicted injury | Infection | Malignancy | Perinatal | SUDIC | Suicide or deliberate self-harm | Trauma |
| Income |  |  |  |  |  |  |  |  |  |  |  |
| Employment | 1.04<br>(1.01-1.07) |  |  |  |  |  |  | 1.04<br>(1.01-1.07) |  | 1.12<br>(1.02-1.23) |  |
| Child Education |  |  |  |  |  | 1.11<br>(1.05-1.18) |  |  |  |  |  |
| Adult Education | 1.03<br>(1.00-1.05) |  | 1.12<br>(1.08-1.16) |  |  |  |  |  |  |  |  |
| Health |  | 1.07 (1.01-1.14) |  | 1.13<br>(1.05-1.21) |  |  |  |  |  |  |  |
| Crime | 0.97<br>(0.95-0.99) |  | 0.95<br>(0.91-0.99) |  |  |  |  |  |  | 0.90<br>(0.82-0.99) |  |
| Geographic Barriers |  |  |  |  |  |  |  |  |  |  |  |
| Wider Barriers | 1.06<br>(1.03-1.08) |  | 1.07<br>(1.02-1.12) |  |  |  |  | 1.06<br>(1.02-1.11) |  |  |  |
| Outdoor Living Environment |  |  | 1.04<br>(1.01-1.07) |  |  |  |  |  |  |  |  |
| Indoor Living Environment | 1.03<br>(1.01-1.05) |  | 1.05<br>(1.01-1.09) |  |  |  |  |  |  |  |  |

\* Adjusted for age, sex, region and rural/urban area
Red boxes show measures where increase in deprivation measures are associated with high risks of death
Green boxes show measures where increase in deprivation measures are associated with lower risks of death

Repeating the main analysis but using the IDACI as the measure of deprivation also gave similar results to the main analysis (unadjusted RR 1.10 (1.09-1.12), p<0.001)); fully adjusted RR 1.08 (1.06-1.09), p<0.001).

## DISCUSSION

Over a fifth of all child deaths may be avoided if the most deprived half of the population had the same mortality as the least deprived, alongside pervasive evidence of a clear gradient of increasing childhood mortality across England as measures of deprivation increase; with a striking finding that this varied little by area, age or other demographic factors. While we acknowledge this gradient is not new[17], the magnitude of the associations is sobering and this study adds detail around the social patterning of potentially modifiable factors. The proportion of modifiable factors increased with increasing deprivation; and this appeared to be restricted to social factors such as financial difficulties, homelessness or poor maternal nutrition. In this detailed analysis an association was seen in most of the categories of death (including the largest category, perinatal); with only causation of death by malignancies, suicide or deliberate self-inflicted harm, and trauma not having clear evidence of an association..

Chance and statistical power are always potential limitations in any statistical analysis, although results in this work were relatively precise. As death notifications are a statutory requirement, the NCMD data is likely to have captured the vast majority of deaths, although some may not have been reported. In addition, postcode data may not have been the child’s only residence; so other influences, unmeasured in this work, may have also impacted on their outcome. However this seems unlikely to have introduced significant bias, and the population nature of the index is more likely to reduce any direct effect of inequalities than introduce a false association. It is important to note that measures of deprivation are derived from neighbourhood measures, and even if directly relevant to the child, assumptions of causality are complex. In contrast, the relative increase in reported modifiable factors, as the index of deprivation increases does suggest that some of the excess mortality estimated here maybe avoidable. This work is novel, with the ability to report and review an individual/record level cohort of childhood mortality, alongside the detailed information obtained at the multi-agency review of every death.

The population attributable risk (of 20%) identifed here is crude, but a worrying estimate of the impact of deprivation in child mortality in England; and would equate to over 700 excess deaths a year in England. It highlights the importance of future work to identify the causal pathways involved and to develop interventions that effectively address the causes and improve survival. While some areas appear relatively unrelated to deprivation (e.g. malignancy) most of these represent relatively uncommon categories of death. Perinatal events, which was the most prevalent, were strongly associated with deprivation and modifiable factors.

We did identify some levels of variation of this association across some measures available to us, but overall the increasing risk with deprivation and child mortality was seen across the whole of England, in all age groups, and communities. Children under 1 living in areas of greater deprivation did appear to have the highest risk of death and this needs further analysis and exploration of potential causal mechanisms but may be due to different disease processes affecting children at different ages, or the differential impact of deprivation at critical periods of the children’s lives. This finding is consistent with the findings from the national perinatal mortality surveillance data, which reported that women living in the most deprived areas are at an 80% higher risk of stillbirth and neonatal death compared to women living in the least deprived areas[18]. Given that death caused by perinatal events also represents the biggest number of childhood deaths in England[19], these findings provide further evidence for the importance of prioritising interventions around pregnancy and the start of life when parents are especially open to support, and targeting families at higher risk[1]. The Marmot review and subsequent reviews recommend that equity be placed at the heart of national decisions about education policy and funding[1]. This study provides further evidence for continued investment in current policies such as the National Healthy Child Programme which are based in the concept of proportionate universalism and designed to address health inequalities for children aged 0-19[20].

Like the wider association with all deaths, the mechanisms are likely to be highly complex, and a combination of the intergenerational impact of poverty on family health and lifestyle choices such as maternal diet and family nutrition[21], parental smoking[22], as well as the environmental impacts of deprivation, such as housing quality, road traffic pollution, and access to health and social care services which create intersectional disadvantage. Further evaluation of community level interventions is needed, for example there is evidence that programmes such as Sure Start reduced the likelihood of hospitalisation among children of primary school age with greater impact on children living in the most deprived areas[23].

Reviewing the components which make up the deprivation index, it should be noted that many of the measures remain very inter-dependent (e.g. income and education) and interpretation should be cautious. Despite universal healthcare, employment was a key association for several of the cause of death categories, and access to care is likely to be an important mediating factor that is amenable to change[24]. A strong association between child mortality and income inequality has been reported amongst the wealthier OECD countries[25] and the UK has among the highest levels of income inequality in Europe.[26] The highest reported measure of income inequality in the UK over the last decade was in the period April 2019 to March 2020[27] and impacts from the COVID pandemic are likely to have worsened this trend. It is notable that Employment, Adult Education, Wider barriers and Indoor Living Environments appear important predictors of child mortality suggesting that adult employment and education opportunities, and access and improvements to housing, may be the most efficient place to target resources in order to reduce these inequalities. This triangulates with qualitative work which identified the lack of cleanliness, unsuitable accommodation (e.g. overcrowding or damp/mould) and financial issues being commonly reported modifiable factors after a child dies.[12] Some component of reverse causality is possible, with households moving to more deprived areas due to family impact of childhood ill health and disability; although children with chronic health conditions may find accessing services or housing/financial support more difficult than others.[12] One other interesting finding was that death by malignancy did not appear strongly associated with any measure of deprivation, and is a childhood condition where outcomes after diagnosis have improved dramatically in recent decades. This supports the view that delivery of healthcare (at least for this condition) does not appear heavily influenced by social inequality. It may be the case that for some of the other categories of death, for example, preterm birth, much of the impact of deprivation relates to the risks of developing the disease/condition in the first place rather than the healthcare delivery afterwards. However further work, looking at differential impact of outcomes after similar clinical presentation may help clarify this. The unexpected association, in the multivariable model, was that of an inverse relationship (compared to the other data) with measures of crime. While it should be noted that before adjusting for other, correlated, measures of deprivation, increasing measures of crime remained associated with increased risk of childhood death; the finding is interesting, and some component measured in the crime metric provides additional and novel information in this area. Currently the child death review data collection form contains a free text area where social deprivation related factors are noted if considered relevant by the CDOP review panel. The form does not include specific and prompting questions for possible factors relating to social deprivation, and improvements in collecting these data in a standardised format would assist in more detailed analysis of future deaths. Any future analyses should explore the information collected about the circumstances of death and modifiable factors in greater detail while analyses following on from this will also need to interpret the results in the context of the economic and social impact of the COVID-19 pandemic.

## Conclusion

There is evidence of a clear gradient of increasing child mortality across England as measures of deprivation increase; with a striking finding that this varied little by area, age or other demographic factor. Over a fifth of all child deaths may be avoided if the most deprived half of the population had the same mortality as the least deprived. Children dying in more deprived areas may have a greater proportion of avoidable deaths, while adult employment and education opportunities, and access and improvements to housing, may be the most efficient place to target resources in order to reduce these inequalities.

## Data Availability

Aggregate data may be available on request to the corresponding author, and subject to approval by HQIP.

## Acknowledgements

We thank all Child Death Overview Panels (CDOPs) who submitted data for the purposes of this report and all child death review professionals for submitting data and providing additional information when requested.

We thank the NCMD team for technical and administrative support.

## Ethics approval and consent to participate

The NCMD legal basis to collect confidential and personal level data under the Common Law Duty of Confidentiality has been established through the Children Act 2004 Sections M - N, Working Together to Safeguard Children 2018 (https://consult.education.gov.uk/child-protection-safeguarding-and-family-law/working-together-to-safeguard-children-revisions-t/supporting_documents/Working%20Together%20to%20Safeguard%20Children.pdf) and associated Child Death Review Statutory & Operational Guidance https://assets.publishing.service.gov.uk/government/uploads/system/uploads/attachment_data/file/859302/child-death-review-statutory-and-operational-guidance-england.pdf).

The NCMD legal basis to collect personal data under the General Data Protection Regulation (GDPR) without consent is defined by GDPR Article 6 (e) Public task and 9 (h) Health or social care (with a basis in law).

In addition we have obtained confirmation of exemption to require additional ethical approval from the Chair of Central Bristol NHS REC.

## Authors Contributions

David O: I declare that I participated in the study concept and design, contributed to acquisition, analysis and interpretation of data, drafting and review of the manuscript and that I have seen and approved the final version.

SS: I declare that I participated in the study design, contributed to data acquisition, linkage, analysis and interpretation of analysis, drafting and review of the manuscript; and that I have seen and approved the final version.

TW: I declare that I participated in the study design, contributed to data acquisition, linkage, analysis and interpretation of data analyses, reviewing the manuscript; and that I have seen and approved the final version.

Dawn O: I declare that I contributed to study design, interpretation of data analysis, reviewing the manuscript; and that I have seen and approved the final version.

JK: I declare that I contributed to study design, interpretation of data analysis, reviewing the manuscript; and that I have seen and approved the final version.

IW: I declare that I contributed to study design, interpretation of data analysis, reviewing the manuscript; and that I have seen and approved the final version.

KL: I declare that I obtained funding for this work, participated in the study concept and design, contributed to data acquisition and interpretation of data, drafting and reviewing the manuscript; and that I have seen and approved the final version.

