## Appendix 1 for "CHILDHOOD DEATHS, DEPRIVATION, AND MODIFIABLE FACTORS: FINDINGS FROM THE NATIONAL CHILD MORTALITY DATABASE"

**Appendix 1. Sub-domains of deprivation (Weight for the overall IMD in brackets).**

**Income Deprivation (22.5%)**

The Income Deprivation Domain measures the proportion of the population in an area experiencing deprivation relating to low income.

**Employment Deprivation (22.5%)**

The Employment Deprivation Domain measures the proportion of the working-age population in an area involuntarily excluded from the labour market; this includes people who are unable to work due to unemployment, sickness or disability, or caring responsibilities**.**

**Education, Skills and Training Deprivation (13.5%)**

The Education, Skills and Training Domain measures the lack of attainment and skills in the local population. The indicators fall into two sub-domains: one relating to children and young people and one relating to adult skills. The Children and Young People Sub-domain measures the attainment of qualifications and associated measures, while the Adult Skills Sub-domain measures the lack of qualifications in the resident working-age adult population.

**Health Deprivation and Disability (13.5%)**

The Health Deprivation and Disability Domain measures the risk of premature death and the impairment of quality of life through poor physical or mental health. The domain measures morbidity, disability and premature mortality but not aspects of behaviour or environment that may be predictive of future health deprivation.

**Crime (9.3%)**

The Crime Domain measures the risk of personal and material victimisation at local level.

**Barriers to Housing and Services (9.3%)**

The Barriers to Housing and Services Domain measures the physical and financial accessibility of housing and local services. The indicators fall into two sub-domains: the Geographical Barriers Sub-domain, which relates to the physical proximity of local services, and the Wider Barriers Sub-domain which includes issues relating to access to housing such as affordability.

**Living Environment Deprivation (9.3%)**

The Living Environment Deprivation Domain measures the quality of the local environment. The indicators fall into two sub-domains. The Indoors Sub-domain measures the quality of housing; while the Outdoors Sub-domain contains measures of air quality and road traffic accidents.
