## Appendix 2 for "CHILDHOOD DEATHS, DEPRIVATION, AND MODIFIABLE FACTORS: FINDINGS FROM THE NATIONAL CHILD MORTALITY DATABASE"

**Appendix 2. Weights of each sub-decile domain towards the total score, and correlations between domains.**

|  | Income | Employment | Health | Crime | Child Education | Adult Education | Geographic Barriers | Wider Barriers | Indoor Living Environment | Outdoor Living Environment |
| --- | --- | --- | --- | --- | --- | --- | --- | --- | --- | --- |
| Income | 1.000 |  |  |  |  |  |  |  |  |  |
| Employment | 0.938 | 1.000 |  |  |  |  |  |  |  |  |
| Health | 0.800 | 0.849 | 1.000 |  |  |  |  |  |  |  |
| Crime | 0.652 | 0.607 | 0.591 | 1.000 |  |  |  |  |  |  |
| Child Education | 0.733 | 0.723 | 0.659 | 0.456 | 1.000 |  |  |  |  |  |
| Adult Education | 0.784 | 0.799 | 0.701 | 0.499 | 0.769 | 1.00 |  |  |  |  |
| Geographic Barriers | -0.443 | -0.380 | -0.367 | -0.464 | -0.228 | -0.251 | 1.000 |  |  |  |
| Wider Barriers | 0.539 | 0.393 | 0.273 | 0.512 | 0.295 | 0.298 | -0.487 | 1.00 |  |  |
| Indoor Living Environment | 0.173 | 0.137 | 0.168 | 0.187 | 0.124 | 0.047 | -0.191 | 0.133 | 1.00 |  |
| Outdoor Living Environment | 0.257 | 0.153 | 0.131 | 0.447 | 0.009 | 0.083 | -0.410 | 0.575 | 0.150 | 1.00 |

Off-diagonal measures are correlation between sub-deciles of the IMD.
